# The potential impact of novel tuberculosis vaccine introduction on economic growth in low- and middle-income countries

**DOI:** 10.1101/2022.11.23.22282690

**Authors:** Allison Portnoy, Jean-Louis Arcand, Rebecca A. Clark, Chathika K. Weerasuriya, Christinah Mukandavire, Roel Bakker, Edith Patouillard, Nebiat Gebreselassie, Matteo Zignol, Mark Jit, Richard G. White, Nicolas A. Menzies

## Abstract

Most individuals who develop tuberculosis (TB) are working-age adults living in low- and middle-income countries (LMICs). The resulting disability and death impacts economic productivity and burdens health systems. New TB vaccine candidates may reduce this burden. In this study, we estimated the impact of introducing novel TB vaccines on GDP growth in 105 LMICs, using a macroeconomic model to simulate country-level GDP trends between 2020 and 2080, in 2020 USD. We compared scenarios for introduction of infant and adolescent/adult vaccine candidates to a no-new-vaccine counterfactual. Compared to the no-new-vaccine counterfactual, both vaccination scenarios produced greater cumulative GDP in the modelled countries over the study period, equivalent to $1.6 (95% uncertainty interval: $0.8–3.0) trillion for the adolescent/adult vaccine, and $0.2 ($0.1–0.4) trillion for the infant vaccine. These GDP gains were substantially lagged relative to the time of vaccine introduction, and concentrated in countries with higher current TB incidence.

## Background

In 2021, an estimated 10.6 million individuals fell ill with tuberculosis (TB), and 1.5 million individuals with TB died.^1^ Developing new safe, affordable, and effective TB vaccines is seen as a necessary step for more rapidly reducing disease incidence and mortality, and their successful development is a central component of the End TB Strategy approved by the World Health Assembly in 2014.^2,3^ While promising vaccine candidates exist, substantial additional resources will be needed to further develop these candidates. To judge whether these investments are justified, it is important to understand the full range and magnitude of benefits that could result from new TB vaccines, and from different perspectives on what constitutes value.

The primary approach for judging the value of health technologies has been through quantifying the additional health that is produced through introducing the technologies. These health benefits can be denominated in disease-specific measures such as the number of infections averted, or through generic measures such as the number of disability-adjusted life years (DALYs) averted, a measure that combines improvements in the length and quality of life and which is used to make comparisons across diseases. A number of studies have assessed the possible health benefits of TB vaccines,^4,5^ projecting potentially large health benefits for vaccine introduction in high-incidence settings. Studies considering the global impact of TB vaccine reduction have estimated up to 40–50 million TB cases could be averted by an effective vaccine by 2050,^6,7^ as well as substantial reductions in TB drug resistance.^8^ Other studies have placed a monetary value on the health benefits generated by TB vaccine introduction, either by reference to the opportunity cost of healthcare spending^9^, or individual willingness-to-pay to reduce health risks.^10^ In addition to these health impacts, studies have quantified the consequences of TB vaccines in terms of reduced income losses due to averted TB,^7,11^ and reductions in catastrophic health expenditures for TB affected households^11^.

In addition to these individual-level outcomes, it is possible that TB vaccine introduction could have consequences at the level of the whole economy. In contrast to most other vaccine-preventable diseases, most individuals who develop TB are working age adults, with 81% of all notified TB cases in 2021 occurring among individuals 15–64 years old.^12^ While TB is less prevalent than some other infectious diseases, individuals who develop the disease experience a long period of illness during which they may not be able to work, followed by a course of treatment that can take 6–24 months to complete. Nationally-representative surveys conducted among TB patients have described substantial income losses due to reduced ability to work, both before a diagnosis is made and during the treatment episode.^1^ For individuals surviving TB, ongoing chronic disability can affect productivity and ability to afford basic needs.^13,14^ Moreover, over 10% of individuals with TB die from the disease, and both these fatal and non-fatal effects will have consequences for labor force size and participation, which could impact rates of economic growth within TB-affected countries.

Earlier studies have estimated the impact of individual diseases and health risks on rates of economic growth.^15-18^ However, the potential gains to economic growth that could be produced by introducing novel TB vaccines have not been previously estimated. In this study, we examined the impact of introducing novel TB vaccines on GDP and GDP growth in 105 LMICs, and how these impacts varied over time and according to different country characteristics.

## Results

### Total economic impact 2028–2080

Across all 105 LMICs considered in this analysis, the projected economic dividend from adolescent/adult vaccine introduction was estimated to be $1618 (95% uncertainty interval: $764–2988) billion over the 2028–2080 period (Table 1). These gains in GDP are equivalent to a 0.033% (0.027–0.039%) increase in total GDP projected for the study period for all modelled countries.

**Table 1.** Gains to gross domestic product (GDP) due to adolescent/adult tuberculosis vaccines across 2028–2080.

| Country grouping | Absolute gains in GDP<br>(billions US\$2020) | Percentage gain in GDP<br>(%) |
| --- | --- | --- |
| All countries | 1618 (764, 2988) | 0.0326% (0.0266%, 0.0388%) |
| High-TB burden<sup>a</sup> | 1587 (756, 2906) | 0.0395% (0.0328%, 0.0467%) |
| High-TB/HIV burden<sup>a</sup> | 1413 (693, 2562) | 0.0854% (0.0727%, 0.1003%) |
| High-MDR/RR-TB burden<sup>a</sup> | 1522 (727, 2777) | 0.0383% (0.0315%, 0.0454%) |
| Income level<sup>b</sup> |  |  |
| Low income | 53.5 (24.4, 103) | 0.0391% (0.0304%, 0.0474%) |
| Lower middle income | 1422 (681, 2594) | 0.0842% (0.0723%, 0.0985%) |
| Upper middle income | 143 (38.3, 303) | 0.0044% (0.0022%, 0.0060%) |
| World region |  |  |
| AFR | 386 (205, 677) | 0.1166% (0.1008%, 0.1338%) |
| AMR | -6.66 (-19.6, -0.63) | -0.0022% (-0.0062%, -0.0001%) |
| EMR | 8.12 (-2.16, 21.0) | 0.0056% (-0.0020%, 0.0120%) |
| EUR | 1.75 (-9.31, 12.5) | 0.0002% (-0.0027%, 0.0018%) |
| SEAR | 981 (459, 1819) | 0.0846% (0.0690%, 0.1024%) |
| WPR | 247 (95.2, 510) | 0.0094% (0.0069%, 0.0119%) |

Note: Values in parentheses represent equal-tailed 95% uncertainty intervals.
<sup>a</sup> High-TB, high-TB/HIV (HIV-associated TB), and high-MDR/RR-TB (multidrug/rifampicin-resistant TB) burden countries as defined by the World Health Organization.
<sup>b</sup> Low income: Gross national income (GNI) per capita of \$1,085 or less; Lower middle income: GNI per capita of \$1,086 to \$4,225; Upper middle income: GNI per capita of \$4,256 to \$13,205 (World Bank 2021).
Note: All countries include 105 low- and middle-income countries analyzed. AFR = African region; AMR = Region of the Americas; EMR = Eastern Mediterranean region; EUR = European region; GDP = gross domestic product; SEAR = Southeast Asian region; WPR = Western Pacific region.

At $207 ($81–405) billion, the infant vaccine showed smaller, but still substantial, impacts on GDP over the 2028–2080 period (Table 2). These gains are equivalent to a 0.004% (0.003– 0.005%) increase in total GDP across all countries projected for the study period. For both vaccine products, average percentage gains in GDP were substantially higher for the subset of countries identified as ‘high-TB burden’ by the WHO.

**Table 2.** Gains to gross domestic product (GDP) due to infant tuberculosis vaccines across 2028–2080.

| Country grouping | Absolute gains in GDP<br>(billions US\$2020) | Percentage gain in GDP<br>(%) |
| --- | --- | --- |
| All countries | 207 (80.6, 405) | 0.0041% (0.0029%, 0.0053%) |
| High-TB burden<sup>a</sup> | 220 (92.7, 416) | 0.0054% (0.0040%, 0.0068%) |
| High-TB/HIV burden<sup>a</sup> | 190 (82.8, 354) | 0.0114% (0.0085%, 0.0147%) |
| High-MDR/RR-TB burden<sup>a</sup> | 212 (89.6, 403) | 0.0053% (0.0039%, 0.0067%) |
| Income level<sup>b</sup> |  | Income level<sup>b</sup> |
| Low income | 3.46 (-0.30, 10.0) | 0.0023% (-0.0004%, 0.0044%) |
| Lower middle income | 187 (78.8, 358) | 0.0110% (0.0081%, 0.0142%) |
| Upper middle income | 16.2 (-1.31, 43.7) | 0.0005% (-0.0001%, 0.0008%) |
| World region |  | World region |
| AFR | 60.3 (25.0, 117) | 0.0180% (0.0120%, 0.0249%) |
| AMR | -3.62 (-6.29, -2.33) | -0.0012% (-0.0021%, -0.0007%) |
| EMR | -2.16 (-5.33, 0.68) | -0.0017% (-0.0045%, 0.0004%) |
| EUR | -2.92 (-5.95, -1.54) | -0.0007% (-0.0017%, -0.0003%) |
| SEAR | 107 (45.2, 211) | 0.0092% (0.0064%, 0.0128%) |
| WPR | 47.8 (15.8, 104) | 0.0018% (0.0011%, 0.0025%) |

Note: Values in parentheses represent equal-tailed 95% uncertainty intervals.
<sup>a</sup> High-TB, high-TB/HIV (HIV-associated TB), and high-MDR/RR-TB (multidrug/rifampicin-resistant TB) burden countries as defined by the World Health Organization.
<sup>b</sup> Low income: Gross national income (GNI) per capita of \$1,085 or less; Lower middle income: GNI per capita of \$1,086 to \$4,225; Upper middle income: GNI per capita of \$4,256 to \$13,205 (World Bank 2021).
Note: All countries include 105 low- and middle-income countries analyzed. AFR = African region; AMR = Region of the Americas; EMR = Eastern Mediterranean region; EUR = European region; GDP = gross domestic product; SEAR = Southeast Asian region; WPR = Western Pacific region.

### Time trends in economic impact

By calendar year, economic benefits were delayed relative to the timing of vaccine introduction, with the greatest economic benefits accruing at the end of the study period (Figure 1). The impact of the adolescent/adult vaccine achieved earlier impacts due to the vaccination of older cohorts where most TB burden lies, whereas the infant vaccine was estimated to produce positive gains over the overall projection period despite only minimal impact prior to 2050.

**Figure 1.**
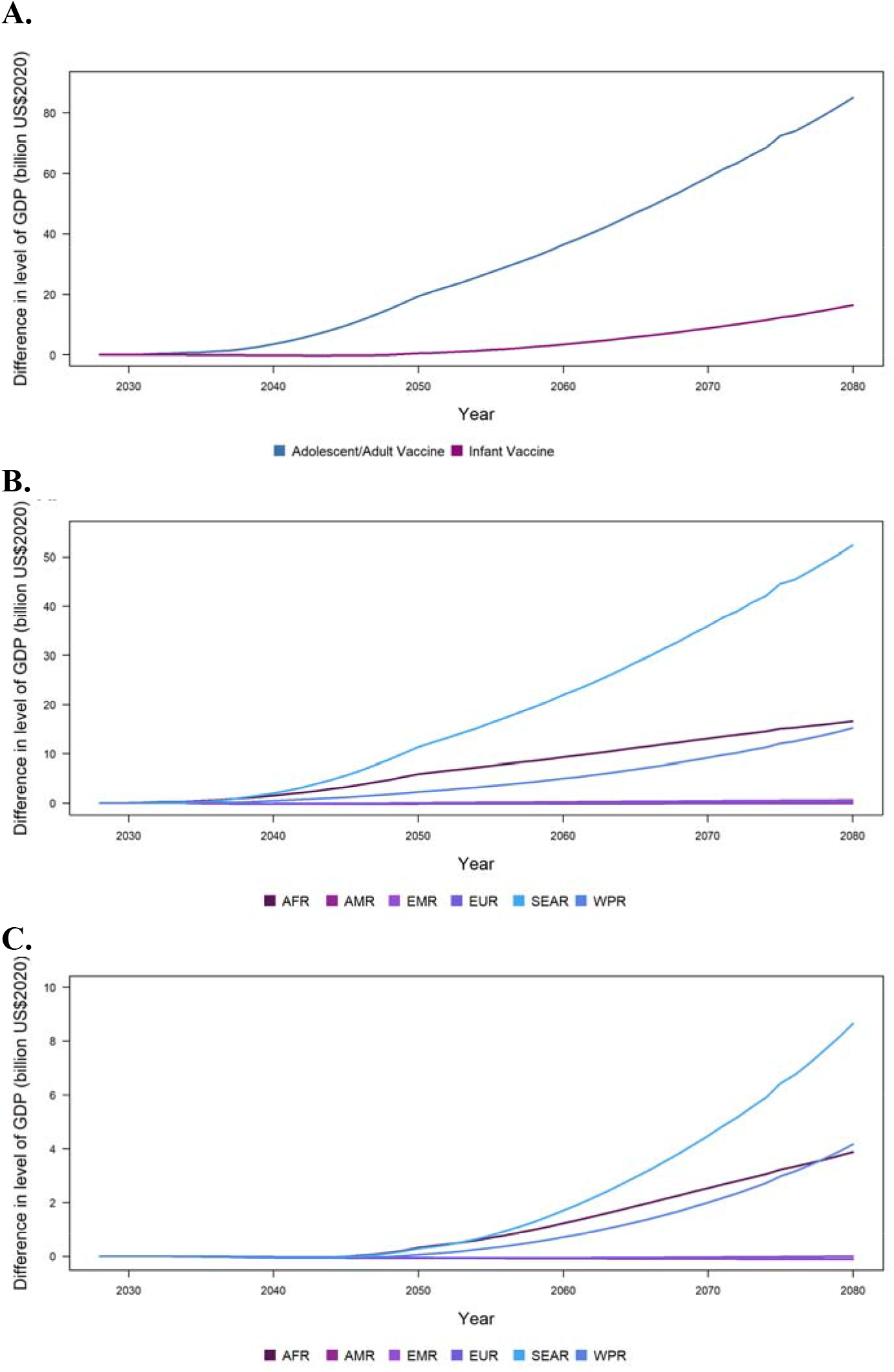
**Time trends in gains of gross domestic product (GDP, in billions US$2020) for all modelled countries, due to adolescent/adult and infant novel tuberculosis vaccination (Panel A); due to adolescent/adult novel tuberculosis vaccine, by world region (Panel B); due to infant tuberculosis vaccine, by world region (Panel C)**.

The WHO regions experiencing the greatest economic growth due to introducing the adolescent/adult and infant novel TB vaccines were the African region (AFR) and Southeast Asian region (SEAR) (Figure 1, Panels B and C). The Eastern Mediterranean region (EMR) and Western Pacific region (WPR) also achieved positive gains to GDP over 2028–2080, whereas the Region of the Americas (AMR) and European region (EUR) with lower TB burden saw positive GDP growth by 2080 but negative gains to GDP over the entire period within the 95% uncertainty intervals. Estimates of the cumulative gains in GDP by decade are given in Appendices S3 and S4.

Figure 2 displays changes in economic growth produced by each vaccine product for successive 5-year periods until 2080, showing the distribution of individual country-level estimates as well as the average impact across countries. For the adolescent/adult vaccine, the impact of vaccine introduction on the rate of economic growth increased progressively until 2046–2050, then declined to over the rest of the analytic period. For the infant vaccine, economic benefits were smaller than estimated for the adolescent/adult vaccine, and substantially lagged relative to the timing of vaccine introduction. The average estimate of additional economic growth was negative for the period 2026–2045, with the opportunity costs of vaccine spending (e.g., deferred educational investments, reduced saving) outweighing the productivity gains resulting from reduced TB incidence. The economic impact of the infant vaccine scenario was positive for all subsequent periods. For both adolescent/adult and infant vaccine scenarios, incremental changes in GDP growth were substantially higher for the 30 countries identified as high-TB-burden by the WHO.

**Figure 2.**
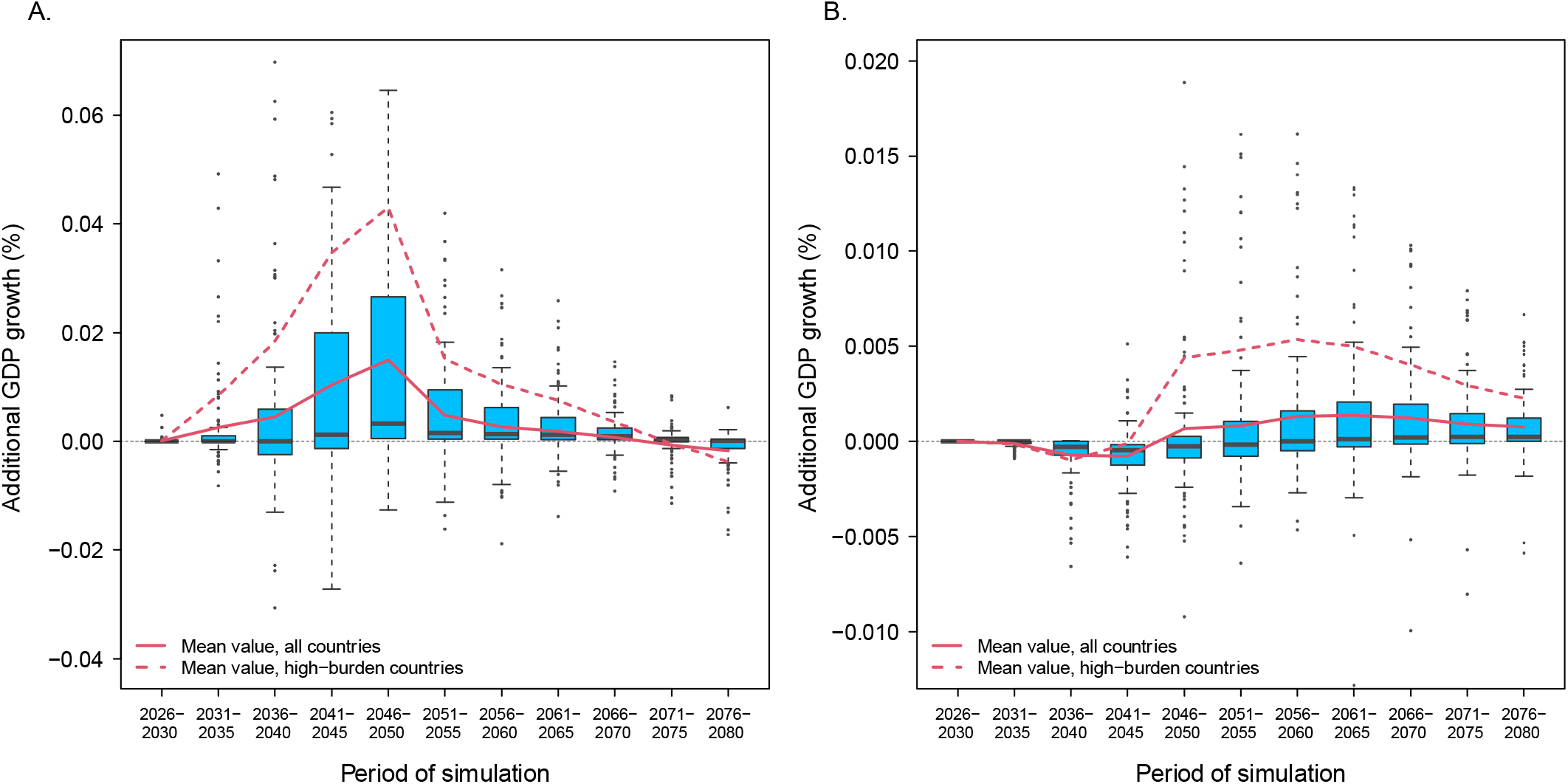
Additional GDP growth in successive 5-year periods from 2026 to 2080 for the adolescent/adult vaccine (Panel A) and infant vaccine (Panel B), as compared to the no-new-vaccine counterfactual. Note: Boxplots represent the distribution of results across individual countries (point estimates), for each 5-year period. Red lines indicate the arithmetic mean of these values for all countries (solid line) and the 30 countries identified as high-TB-burden by the WHO (dashed line). Red shaded regions report 95% uncertainty intervals. Note different y-axis scales.

### Relationship between economic impact and country characteristics

Table 3 reports the results of linear regression models describing the relationship between the percentage gains in GDP produced by each vaccine scenario (percentage increase in GDP over the 2028–2080 period) and selected country characteristics. In these analyses, higher current TB incidence (p<0.001) and earlier vaccine introduction (p<0.001) were associated with greater economic impact for both vaccine scenarios, controlling for other factors. Current TB incidence showed the strongest relationship with economic impact, with a one standard deviation increase in TB incidence associated with a 0.050% (0.040–0.059%) increase in GDP for the adolescent/adult vaccine scenario, and a 0.007% (0.005–0.100%) increase in GDP for the infant vaccine scenario. Figure 3 shows the relationship between the additional economic growth produced by the adolescent/adult vaccine and current TB incidence rate and vaccine introduction year. Of the macroeconomic variables, only the elasticity of physical capital had a statistically significant relationship with the impact of vaccine introduction, with higher vaccine impact estimated for countries with lower values for this parameter, controlling for other predictors (Table 3).

**Table 3.**
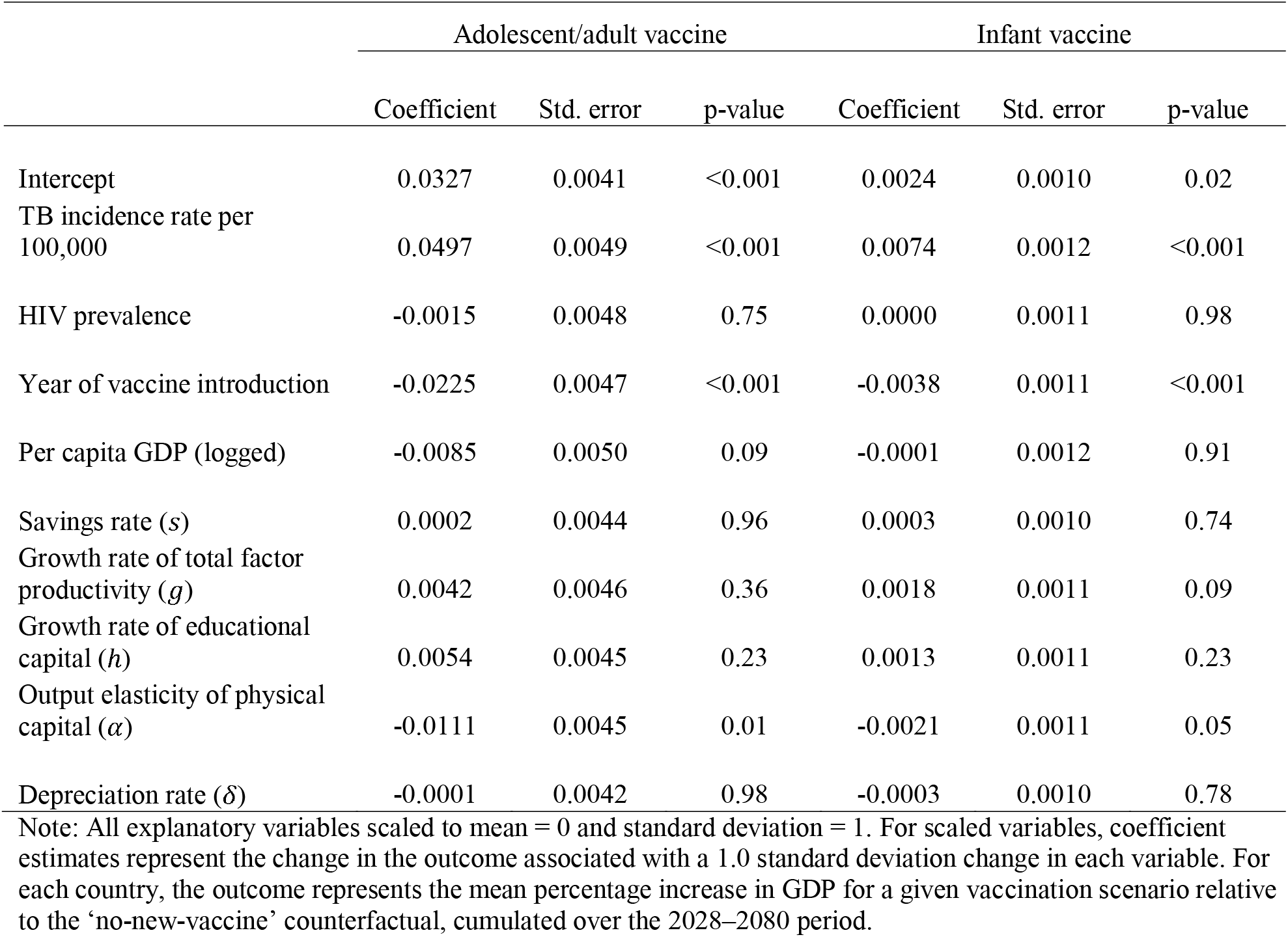
Relationship between additional GDP growth in each vaccine scenario (relative to the ‘no-new-vaccine’ counterfactual), and selected country characteristics.

**Figure 3.**
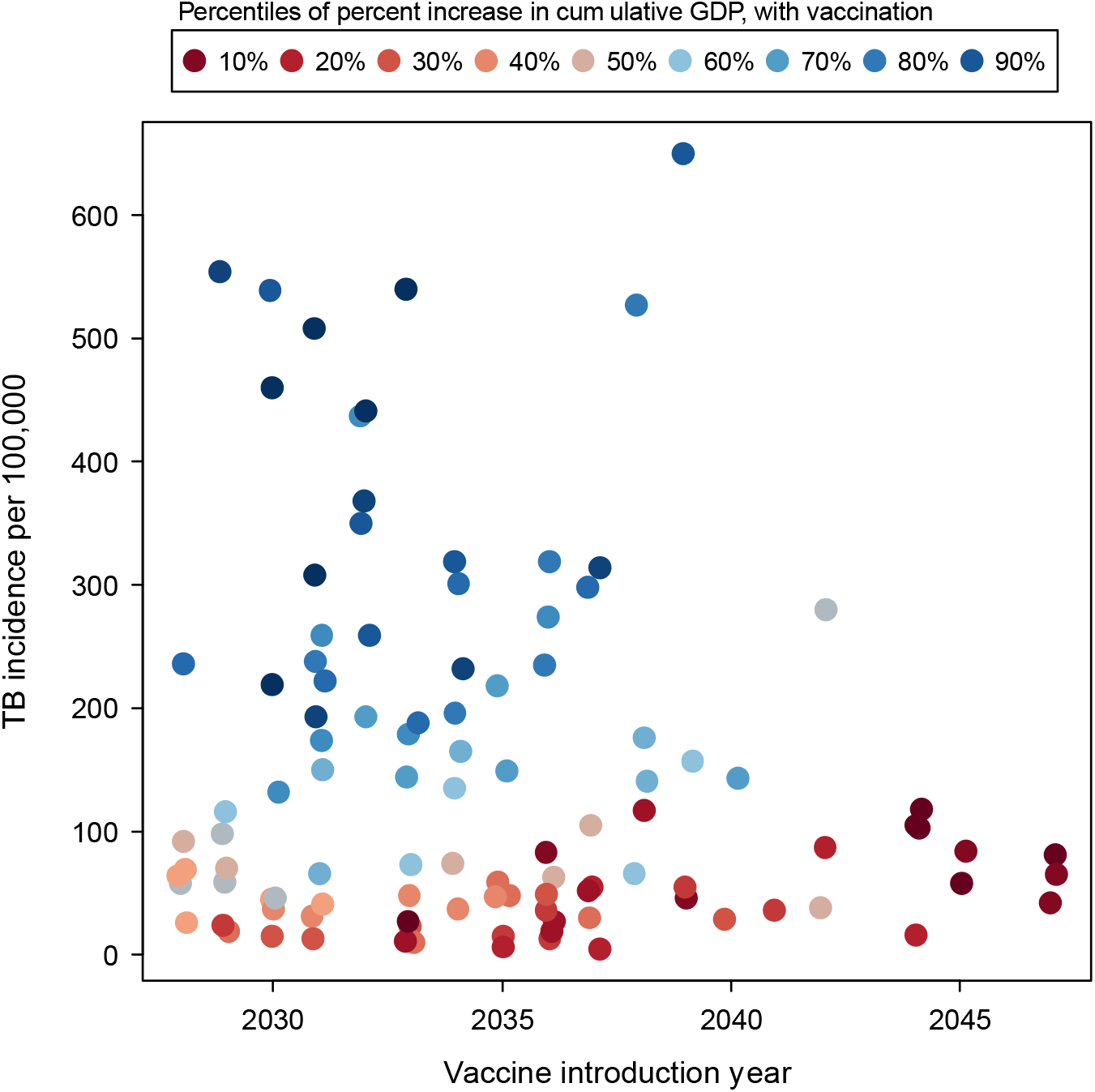
Relationship between additional GDP growth produced by the adolescent/adult vaccine over 2028–2080, current TB incidence level, and vaccine introduction year. Note: Each point represents point-estimate results for an individual country. Color scale ranges from low percentage increase in GDP relative to other countries (reds) to high percentage increase in GDP relative to other countries (blues). Vaccine introduction year values jittered to display overlapping points.

### Sensitivity analyses

Compared to the main analysis (estimated GDP gains of $1687 ($816–3076) billion for the adolescent/adult vaccine, and $219 ($91–422) billion for the infant vaccine) the first alternative specification excluding patient-incurred costs resulted in estimated GDP gains that were 8–10% smaller (Appendices S5 and S6 for adolescent/adult and infant vaccines respectively) than estimated for the base-case specification. Under the second alternative specification, excluding costs incurred by domestic governments resulted in estimated GDP gains that were 5–17% greater than estimated for the base-case specification (Appendices S7 and S8). Under the third alternative specification, including costs attributed to international donors resulted in estimated GDP gains that were 10–22% lower than estimated for the base-case specification (Appendices S9 and S10).

Low-growth and high-growth specifications had a greater impact on the estimated economic impact of vaccine introduction. For the low-growth scenario, estimated GDP gains were 37–43% lower than estimated for the base-case specification (Appendices S11 and S12). Under the high-growth scenario, estimated GDP gains were 36–40% higher than estimated for the base-case specification (Appendices S13 and S14).

Under the final alternative specification, with the economic impact of non-fatal health losses values represented as income losses rather than reductions in the labor supply, estimated GDP gains were 15–21% lower than estimated for the base-case specification (Appendices S15 and S16).

## Discussion

In this study we estimated changes in the economic performance of 105 LMICs that would result from the introduction of novel TB vaccines, based on characteristics specified in the WHO PPCs,^19^ and promising evidence from ongoing clinical trials.^20,21^ At a global level, both vaccine scenarios were shown to produce greater cumulative GDP in the modelled countries over the analytic period, with US$1618 billion in economic gains estimated for the adolescent/adult vaccine, and $207 billion for the infant vaccine. For both vaccine products, economic benefits were concentrated in countries with a high burden of TB, and lagged relative to the timing of vaccine introduction, particularly for the infant vaccine. The magnitude of GDP gains was strongly associated with higher country TB incidence level, and with earlier vaccine introduction.

Study results were robust to most alternative analytic specifications. The sensitivity analyses with the greatest impact involved changes in economic growth projections, with high and low growth projections leading to estimates of vaccine impact that were substantially larger and smaller, respectively, than estimated in the main analysis, illustrating the sensitivity of results to future economic conditions. Another sensitivity analysis that led to larger changes in the results was where we modified the approach taken to representing the macroeconomic impact of non-fatal illness. In this comparison, modelling non-fatal illness as directly reducing the labor supply (main analysis) produced economic impacts that were one-fifth greater than when we modelled non-fatal illness as producing additional costs for TB-affected households. Despite these differences, all the alternative analytic specifications suggested the economic gains of vaccine introduction would be substantial.

It is important to note that the economic impacts estimated in this study would not be realized as an explicitly monetary gain, but instead as small, widely-spread improvements in living standards within TB-affected countries. While small relative to overall changes in GDP, these economic gains are large relative to the investments needed to successfully develop TB vaccines. For example, the 2023–2030 Stop TB Global Plan includes $10 billion in research funding to successfully develop TB vaccines, less than 1% of the absolute GDP gains estimated for the adult vaccine scenario.^22^ Of course, the economic benefits of TB vaccine introduction would be substantially lagged relative to the timing of investments needed to develop these vaccines, which is a feature of vaccine interventions more generally. For this reason, decisions to invest in TB vaccines based on macroeconomic impacts must consider the longer time frame over which benefits will accrue, as well as the fact that these benefits will be largely invisible, despite their magnitude.

A strength of this analysis is the approach used to estimated economic outcomes. Most studies estimating the economic impact of disease adopt a ‘Cost of Illness’ approach, in which incidence of a given condition is multiplied by the range of per-episode costs borne by patients, governments, and broader society, in order to sum the total costs resulting from the disease. However, this ‘Cost of Illness’ approach does not take into account the dynamic relationships between labor supply, capital accumulation, and economic growth. The macroeconomic model used for this analysis allows for these relationships. Moreover, for this analysis, we also extended this model to incorporate the effects of morbidity on individual productivity, a limitation noted in earlier applications.^23^

This analysis has several limitations. Firstly, the macroeconomic model relies on projections of GDP that are inherently uncertain. Unpredictable natural disasters or conflicts could alter these projections and affect the conclusions drawn by this analysis. However, we attempted to address this limitation by examining both high- and low-growth scenarios for GDP, and allowing for stochastic changes in macroeconomic variables based on past performance. Secondly, we assumed a working age population of ages 15–69 contributing to the labor market. This assumption excludes older workers as well as informal caregivers that may affect labor force participation. Thirdly, we based the characteristics of new TB vaccines in our analyzed scenarios on the WHO PPCs,^19^ but a final product may differ in terms of effectiveness or duration of protection.^1^ Finally, we assumed that TB trends in the no-new-vaccine baseline would follow their historical trajectory. If there were aggressive scale-up of non-vaccine interventions (as envisaged by recent global TB strategy), this would reduce the incremental impact of a new vaccine.^1^

To guide global efforts to accelerate TB control, it is critical to understand the full range of consequences of possible intervention options. If shown to be effective, TB vaccines promise to protect individuals from the risks posed by TB infection, leading to population-level reductions in TB burden,^6^ and TB-related catastrophic costs.^11^ This study demonstrates that, under a range of assumptions, novel TB vaccines could have a further beneficial impact for TB-affected LMICs, through increasing rates of economic growth. These impacts should be viewed not as alternative way of quantifying the benefits of vaccination, but as an additional benefit, which can be added to the value of the health gains and financial risk protections produced through reduced TB morbidity and mortality. This full range of outcomes should be considered when weighing the value of further investment in TB vaccine development.

## Methods

### Analytic approach

We used a mathematical model of *Mycobacterium tuberculosis* (*Mtb*) transmission, progression, care and prevention to simulate changes in population health outcomes that would be produced by introduction of a new TB vaccine (as compared to a ‘no-new-vaccine’ counterfactual) in each of 105 LMICs, and a related costing model that simulated changes in health service costs (TB vaccination, TB diagnosis and treatment, HIV treatment) affected by vaccine introduction.

Detailed methods and outcomes of these models are described elsewhere.^6,9^ We used the results of these analyses to generate inputs for an existing macroeconomic model, which we used to translate changes in disease-related mortality, morbidity, and health service utilization into projected macroeconomic outcomes.

### Analytic scenarios

We simulated the macroeconomic impact of TB vaccine introduction for two separate vaccine products, both assumed to prevent disease with a 10-year average duration of protection. These characteristics were based on WHO Preferred Product Characteristics (PPCs) for novel TB vaccines.^19^ The infant TB vaccine was assumed to be effective only among individuals without prior *Mtb* infection with 80% efficacy, and would be introduced as part of the routine infant vaccine schedule in each country. The adolescent/adult TB vaccine was assumed to be effective for both infected and uninfected individuals with 50% efficacy, and would be provided through an initial mass vaccination campaign of all age-groups followed by routine vaccination of 9-year-olds. We examined scenarios for each vaccine product individually, and assumed vaccination would be introduced and scaled-up over a 5-year period. The vaccine introduction year varied by country (based on an analysis of factors affecting new vaccine adoption), ranging from 2028 and 2047.^6^ We compared these scenarios to a counterfactual ‘no-new-vaccine’ scenario in order to calculate the incremental impact of vaccine introduction on country GDP. In all scenarios we assumed that the quality of and access to all health services (apart from the new TB vaccination) would be maintained at current levels.

In the base-case analysis, in addition to vaccination program costs, we included costs from the societal perspective for direct medical costs TB/HIV treatment (both government- and patient-level costs), and patient-level direct non-medical costs (additional details on cost assumptions are described elsewhere^9^). For all unit cost inputs related to service provision, we subtracted the share contributed by international donors, to obtain unit costs reflecting the contribution of domestic resources to service provision. For vaccination costs, we assumed a donor share stratified by World Bank income level (69.4% for low-income countries, 39.4% for lower middle-income countries, 0.2% for upper middle-income countries) based on Ikilezi, et al.^24^ For TB diagnosis and treatment costs, we estimated country-specific donor shares using spending estimates from the WHO Global TB Report 2022 (Appendix S1).^1^ For antiretroviral therapy costs, we assumed a country-specific donor share based on spending estimates from the Global Burden of Disease Collaborative Network at IHME (Appendix S1).^25^ All costs underlying the macroeconomic analysis were calculated in 2020 constant US dollars.

We simulated macroeconomic outcomes over 2028–2080, to capture the long-term consequences of vaccine introduction. For the period during and immediately following vaccine introduction (2028–2050) we simulated model inputs using the *Mtb* transmission and costing models described earlier. For long-term outcomes (2050–2080) we extrapolated population health inputs by applying UN Population Division mortality rate projections by age, and year to the population of each country at the end of 2050 for each analytic scenario.^26^ Cost inputs for 2050–2080 were assumed to change in proportion to population growth (based on 2050 values) for each scenario.

### Macroeconomic model

The WHO EPIC (Economic Projections of Illness and Cost) model is based on a standard human capital-augmented Solow model of economic growth, and has been used in prior studies to project the macroeconomic impact of various health conditions.^23,27-31^ We adapted this model to estimate the effect of vaccine introduction on the economic output of each modelled country, as produced by averted TB morbidity and mortality and related changes in health service utilization. Full details of model specification are provided in Supplementary Materials (Appendix S2). Changes in TB mortality were assumed to influence macroeconomic outcomes through changes in the labor supply, with the size and age-structure of the available workforce (stock of individuals aged 15–69, adjusted for labor force participation and labor quality) modified to reflect the additional survival estimated for the TB vaccination scenarios. We assumed that labor force participation rates and labor quality within each single-year age group were the same across vaccine and no-new-vaccine scenarios. To account for changes in non-fatal health outcomes, we subtracted from this labor supply the Years Lived with Disability (YLDs) estimated for the vaccine introduction scenarios as compared to the no-new-vaccine scenario, for each year of age. These YLDs represent the incremental person-time spent in ill-health, scaled by the relative disability associated with a given condition.^32^ We did not account for any additional losses of productivity within the household to care for sick individuals, but varied this in sensitivity analysis. Changes in domestic health spending (total costs of modelled health services net of the international donor share for these services) were subtracted directly from the total stock of physical capital. In sensitivity analysis we examined alternative assumptions for the macroeconomic impact of changes in healthcare spending. Other variables determining macroeconomic outcomes (savings rate, growth rate of total factor productivity, growth rate of educational capital, output elasticity of physical capital, and depreciation rate for each country) were derived from published sources (Appendix S2) and assumed to be unaffected by vaccine introduction. With these specifications, we used the EPIC model to project total annual GDP for each modelled country over 2020–2080.

### Outcomes

We estimated incremental changes in GDP within each country from vaccine introduction until 2080, by comparing each vaccine scenario to the no-new-vaccine counterfactual. We summarized results as absolute and percentage differences in GDP, changes in per-capita GDP, and time trends in the rate of GDP growth within modelled countries. We report results according to major country groupings (global, WHO high-TB burden grouping,^33^ World Bank income level,^34^ and WHO region). We also estimated linear regression models for the percentage change in GDP under vaccination scenarios as a function of selected country characteristics (current TB incidence level, HIV incidence level, country per-capita GDP, year of vaccine introduction) and country-specific macroeconomic variables (mean values for the savings rate, growth rate of total factor productivity, growth rate of educational capital, output elasticity of physical capital, and depreciation rate), to describe the relationship between these factors and the economic gains produced by TB vaccination. Predictors were standardized (re-scaled to mean = 0.0 and standard deviation = 1.0) prior to fitting these models, to facilitate comparisons. With these standardized predictors, regression coefficients represent the estimated change in GDP growth associated with a one standard deviation change in each predictor.

### Statistical analysis

Uncertainty intervals (95% coverage, equal-tailed) for the projected gains in GDP were generated via a 2^nd^-order Monte Carlo simulation. To implement this, we derived 1000 simulated trajectories of epidemiological and intervention cost outcomes from the *Mtb* transmission and costing models. For country-specific values for the macroeconomic parameters we generated future values for each country and year using a semi-parametric bootstrapping approach. First, we fit random-effects regression models to estimate country-average values for each parameter, based on recent data (2008–2019) reported for each country. Any missing values were assumed to be missing-at-random conditional on WHO world region and country income level. Second, we resampled with replacement from the residuals of these regression models and combined these with the country-average values, producing a time series of future values for each parameter and country. We combined all sources of uncertainty to generate 1000 estimated trajectories of annual GDP for each modelled scenario and country. We took the mean of these 1000 simulations to produce point estimate results for each outcome of interest, and calculated 95% intervals as the 2.5^th^ and 97.5^th^ percentiles of the distribution of results for each outcome of interest.

### Sensitivity analysis

Compared to the base-case cost assumptions (including TB-related health services costs incurred by patients and domestic government, excluding costs borne by international donors), we examined three alternative approaches to the specification of health services costs in the macroeconomic analysis: (1) modifying the base-case to exclude patient-incurred costs, under the assumption that estimated changes in household healthcare spending would be offset by matching changes in other consumption; (2) modifying the base-case to exclude costs borne by domestic government, under the assumption that estimated changes in government spending would be offset by matching changes in taxation; and (3) modifying the base-case to include costs borne by international donors, assuming that the absolute value of donor spending in each country would not change between vaccination and no-new-vaccine scenarios, such that domestic governments would absorb all incremental health service costs.

We also examined how results would change under low and high economic growth trajectories. We operationalized the low-growth specification by estimating results from the 20% of simulations for each country that had the lowest average annual growth over the 2020–2080 period under the no-new-vaccine scenario. Similarly, the high-growth specification was estimated from the 20% of simulations for each country with the highest average annual growth over 2020–2080 period under the no-new-vaccine scenario.

Finally, we estimated results with an alternative approach for incorporating the impact of TB-related morbidity. Compared to the main analysis, in which non-fatal illness was assumed to reduce the labor supply (with sick individuals unable to participate in the workforce), this specification instead quantified these changes through the income losses experienced by TB-affected households due to reduced productivity. These values were based on data from nationally-representative TB patient surveys.^35^ These income losses were values in monetary units, and added to other costs borne by patients.

## Data Availability

The data that support the findings of this study are available from the corresponding author upon reasonable request.

## Acknowledgements

We are grateful for the support of the World Health Organization for funding this research (2020/985800-0). We thank all the attendees at the WHO meetings on the Full Value Assessment of TB Vaccines for insightful advice and direction.

## Author contributions

AP, JLA, and NAM conceptualized and designed the study. AP and JLA conducted the analysis with support from NAM, RAC, and RGW. AP, JLA, and NAM drafted the initial manuscript and approved the final manuscript as submitted. RAC, CKW, CM, RB, EP, NG, MZ, MJ, and RGW critically reviewed the analysis, reviewed and revised the manuscript, and approved the final manuscript as submitted. All authors read and approved the final manuscript.

## Competing interests

RAC is funded by BMGF (INV-001754) and received a grant from the Canadian Centennial Scholarship Fund. CKW is funded by UKRI/MRC (MR/N013638/1). RGW is funded by the Wellcome Trust (218261/Z/19/Z), NIH (1R01AI147321-01), EDTCP (RIA208D-2505B), UK MRC (CCF17-7779 via SET Bloomsbury), ESRC (ES/P008011/1), BMGF (OPP1084276, OPP1135288 & INV-001754), and the WHO (2020/985800-0). Members of the funder participated as authors on the study. All other authors declare no conflicts of interest.

**Correspondence** and requests for materials should be addressed to Allison Portnoy.

