## Supplementary Materials for "The potential impact of novel tuberculosis vaccine introduction on economic growth in low- and middle-income countries"

### Appendix S1. International donor share of TB and HIV/AIDS spending based on development assistance for health spending estimates.

| **Country** | **WHO Region** | **Income level^a^** | **Vaccine introduction year** | **TB international donor share^1^** | **HIV/AIDS international donor share^2^** |
| --- | --- | --- | --- | --- | --- |
| Afghanistan | EMR | LIC | 2031 | 0.79 | 0.79 |
| Angola | AFR | LMIC | 2032 | 0.11 | 0.24 |
| Albania | EUR | UMIC | 2035 | 0.00 | 0.02 |
| Argentina | AMR | UMIC | 2031 | 0.00 | 0.00 |
| Armenia | EUR | UMIC | 2033 | 0.25 | 0.49 |
| Azerbaijan | EUR | UMIC | 2028 | 0.21 | 0.34 |
| Burundi | AFR | LIC | 2044 | 0.45 | 0.94 |
| Benin | AFR | LMIC | 2037 | 0.71 | 0.74 |
| Burkina Faso | AFR | LIC | 2039 | 0.73 | 0.45 |
| Bangladesh | SEAR | LMIC | 2035 | 0.74 | 0.48 |
| Bulgaria | EUR | UMIC | 2029 | 0.05 | 0.04 |
| Belarus | EUR | UMIC | 2028 | 0.03 | 0.21 |
| Bolivia | AMR | LMIC | 2037 | 0.28 | 0.17 |
| Brazil | AMR | UMIC | 2030 | 0.00 | 0.00 |
| Bhutan | SEAR | LMIC | 2034 | 0.28 | 0.29 |
| Botswana | AFR | UMIC | 2028 | 0.36 | 0.24 |
| Central African Republic | AFR | LIC | 2033 | 0.51 | 0.90 |
| China | WPR | UMIC | 2029 | 0.01 | 0.00 |
| Côte d'Ivoire | AFR | LMIC | 2034 | 0.43 | 0.87 |
| Cameroon | AFR | LMIC | 2031 | 0.56 | 0.70 |
| Colombia | AMR | UMIC | 2030 | 0.01 | 0.02 |
| Costa Rica | AMR | UMIC | 2033 | 0.00 | 0.04 |
| Cuba | AMR | UMIC | 2035 | 0.00 | 0.06 |
| Dominican Republic | AMR | UMIC | 2031 | 0.09 | 0.36 |
| Ecuador | AMR | UMIC | 2033 | 0.00 | 0.08 |
| Egypt | EMR | LMIC | 2033 | 0.07 | 0.01 |
| Eritrea | AFR | LIC | 2047 | 0.94 | 0.87 |
| Ethiopia | AFR | LIC | 2030 | 0.47 | 0.87 |
| Fiji | WPR | UMIC | 2031 | 0.20 | 0.01 |
| Gabon | AFR | UMIC | 2038 | 0.28 | 0.04 |
| Georgia | EUR | UMIC | 2029 | 0.48 | 0.38 |
| Ghana | AFR | LMIC | 2040 | 0.59 | 0.51 |
| Guinea | AFR | LIC | 2033 | 0.71 | 0.54 |
| Gambia | AFR | LIC | 2039 | 0.52 | 0.88 |
| Equatorial Guinea | AFR | UMIC | 2042 | 0.03 | 0.01 |
| Guatemala | AMR | UMIC | 2036 | 0.23 | 0.21 |
| Honduras | AMR | LMIC | 2037 | 0.33 | 0.18 |
| Indonesia | SEAR | UMIC | 2034 | 0.36 | 0.32 |
| India | SEAR | LMIC | 2033 | 0.24 | 0.15 |
| Iran | EMR | UMIC | 2031 | 0.06 | 0.03 |
| Iraq | EMR | UMIC | 2033 | 0.07 | 0.01 |
| Jordan | EMR | UMIC | 2037 | 0.15 | 0.35 |
| Kazakhstan | EUR | UMIC | 2028 | 0.06 | 0.14 |
| Kenya | AFR | LMIC | 2032 | 0.39 | 0.50 |
| Kyrgyz Republic | EUR | LMIC | 2044 | 0.31 | 0.59 |
| Cambodia | WPR | LMIC | 2036 | 0.29 | 0.76 |
| Lao People’s Democratic Republic | WPR | LMIC | 2035 | 0.49 | 0.71 |
| Liberia | AFR | LIC | 2037 | 0.98 | 0.58 |
| Libya | EMR | UMIC | 2035 | 0.00 | 0.01 |
| Sri Lanka | SEAR | LMIC | 2028 | 0.15 | 0.15 |
| Lesotho | AFR | LMIC | 2039 | 0.66 | 0.62 |
| Morocco | EMR | LMIC | 2029 | 0.05 | 0.28 |
| Moldova, Republic of | EUR | LMIC | 2034 | 0.14 | 0.59 |
| Madagascar | AFR | LIC | 2031 | 0.62 | 0.69 |
| Maldives | SEAR | UMIC | 2034 | 0.00 | 0.01 |
| Mexico | AMR | UMIC | 2029 | 0.00 | 0.00 |
| Mali | AFR | LIC | 2037 | 0.46 | 0.77 |
| Myanmar | SEAR | LMIC | 2031 | 0.88 | 0.77 |
| Montenegro | EUR | UMIC | 2044 | 0.05 | 0.00 |
| Mongolia | WPR | LMIC | 2032 | 0.15 | 0.44 |
| Mozambique | AFR | LIC | 2032 | 0.72 | 0.92 |
| Mauritania | AFR | LMIC | 2042 | 0.77 | 0.10 |
| Malawi | AFR | LIC | 2038 | 0.91 | 0.85 |
| Malaysia | WPR | UMIC | 2028 | 0.00 | 0.02 |
| Namibia | AFR | UMIC | 2030 | 0.14 | 0.31 |
| Niger | AFR | LIC | 2036 | 0.82 | 0.68 |
| Nigeria | AFR | LMIC | 2030 | 0.63 | 0.63 |
| Nicaragua | AMR | LMIC | 2047 | 0.25 | 0.40 |
| Nepal | SEAR | LMIC | 2036 | 0.35 | 0.31 |
| Pakistan | EMR | LMIC | 2031 | 0.80 | 0.60 |
| Peru | AMR | UMIC | 2029 | 0.03 | 0.01 |
| Philippines | WPR | LMIC | 2030 | 0.36 | 0.13 |
| Papua New Guinea | WPR | LMIC | 2032 | 0.56 | 0.58 |
| Paraguay | AMR | UMIC | 2035 | 0.17 | 0.14 |
| Russian Federation | EUR | UMIC | 2030 | 0.00 | 0.02 |
| Rwanda | AFR | LIC | 2045 | 0.78 | 0.82 |
| Sudan | EMR | LIC | 2036 | 0.87 | 0.68 |
| Senegal | AFR | LMIC | 2038 | 0.39 | 0.74 |
| Solomon Islands | WPR | LMIC | 2047 | 0.44 | 0.07 |
| Sierra Leone | AFR | LIC | 2037 | 0.22 | 0.97 |
| El Salvador | AMR | LMIC | 2039 | 0.29 | 0.15 |
| Serbia | EUR | UMIC | 2036 | 0.00 | 0.01 |
| South Sudan | AFR | LIC | 2034 | 1.00 | 0.98 |
| São Tomé and Principe | AFR | LMIC | 2044 | 0.66 | 0.76 |
| Suriname | AMR | UMIC | 2040 | 0.36 | 0.20 |
| Swaziland | AFR | LMIC | 2036 | 0.79 | 0.62 |
| Syrian Arab Republic | EMR | LIC | 2036 | 0.99 | 0.45 |
| Chad | AFR | LIC | 2033 | 0.48 | 0.68 |
| Togo | AFR | LIC | 2041 | 0.55 | 0.69 |
| Thailand | SEAR | UMIC | 2031 | 0.14 | 0.03 |
| Tajikistan | EUR | LIC | 2045 | 0.36 | 0.68 |
| Timor-Leste | SEAR | LMIC | 2031 | 0.77 | 0.42 |
| Tunisia | EMR | LMIC | 2036 | 0.00 | 0.24 |
| Turkey | EUR | UMIC | 2030 | 0.00 | 0.00 |
| Tanzania, United Republic of | AFR | LMIC | 2031 | 0.69 | 0.89 |
| Uganda | AFR | LIC | 2034 | 0.87 | 0.83 |
| Ukraine | EUR | LMIC | 2033 | 0.24 | 0.38 |
| Uzbekistan | EUR | LMIC | 2038 | 0.36 | 0.47 |
| Venezuela | AMR | UMIC | 2035 | 0.30 | 0.00 |
| Vietnam | WPR | LMIC | 2038 | 0.39 | 0.44 |
| Vanuatu | WPR | LMIC | 2042 | 0.45 | 0.16 |
| Yemen | EMR | LIC | 2036 | 0.87 | 0.34 |
| South Africa | AFR | UMIC | 2029 | 0.12 | 0.20 |
| Zambia | AFR | LIC | 2034 | 0.55 | 0.86 |
| Zimbabwe | AFR | LMIC | 2032 | 0.83 | 0.70 |

^a^ LIC: Gross national income (GNI) per capita of $1,085 or less; LMIC: GNI per capita of $1,086 to $4,225; UMIC: GNI per capita of $4,256 to $13,205 (World Bank 2021).

Note: All countries include 105 low- and middle-income countries analyzed. AFR = African region; AMR = Region of the Americas; EMR = Eastern Mediterranean region; EUR = European region; LIC = low-income; LMIC = lower middle-income; SEAR = Southeast Asian region; TB = tuberculosis; UMIC = upper middle-income; WPR = Western Pacific region.

### Appendix S2. Technical specifications of the macroeconomic model

The WHO EPIC (Economic Projections of Illness and Cost) model^3-8^ model is based on a standard human capital augmented Solow growth model, with a linear-homogeneous Cobb-Douglas production function:

| $Y_{it}=\gamma_{i}A_{it}*K_{it}^{\alpha_{it}}*(H_{it}*L_{it})^{1-\alpha_{it}}$ where $0<\alpha<1$, | (1) |
| --- | --- |

where $Y_{it}$ is the GDP of country $i$ in year $t$, $K$ is physical capital, $A_{it}$ is total factor productivity, $L_{it}$ is the aggregate labor force (additional details below), and $H_{it}$ is an educational capital index, i.e., returns to education that increase labor quality. The product $H_{it}*L_{it}$ therefore represents the stock of aggregate labor augmented by education. Total factor productivity $A_{it}$ (which here is Hicks-neutral) enters multiplicatively in the production function as an increasing scale factor. Total factor productivity is an estimate of the change in economic growth that occurs due to factors other than changes in the labor force or capital stock (e.g., technological advancement). The elasticity of output with respect to physical capital is denoted by $\alpha_{it}\in(0, 1)$ and the elasticity of output with respect to labor is denoted by ${(1-\alpha}_{it})\in(0, 1)$. The scaling factor $\gamma_{i}$ is a calibration parameter used to fit the model to reported GDP values (indicator *NY.GDP.MKTP.K* from the World Bank’s World Development Indicators) so that the two are identical in the first year of the projection. For most countries, the initial year used for calibration was 2020. Exceptions, when 2020 data on GDP were not available, were: ERI (2011), SSD (2015) and SYR (2018).

The law of motion for physical capital is given by the equation:

$K_{it}=s_{it}Y_{i(t-1)}+(1-\delta_{it})K_{i(t-1)}$,

(2)

where $s_{it}$ is the savings rate, which was assumed to be exogenous, as well as country- and year-specific. The depreciation rate $\delta_{it}$ was also assumed to be country- and year-specific.

We extracted estimates for the savings rate $s_{it}$ from the World Bank’s World Development Indicators (indicator: *NE.GDI.TOTL.ZS*).^9^ We extracted estimates for the depreciation rate $\delta_{it}$ from the latest Penn World Tables (indicator: *delta*).^10^ Under the assumption of competitive markets, the elasticity of output with respect to physical capital $\alpha_{it}$ is equal to labor income as a share of GDP. We derived values for this variable from ILOSTAT (indicator: *LAP_2GDP_NOC_RT_A*).^11^

The labor force was defined as the stock of individuals of working age (ages 15–69) who participate in the labor market, with data organized by single-year age cohorts. Historical estimates (up to 2020) and future projections (to 2080) of demographic inputs were obtained from the UN Population Division (World Population Prospects 2019^12^), with $N_{ait}$ denoting the population of age group $a$ in country $i$ in year $t$.

The raw stock of labor for each age group was calculated by multiplying the base population by the labor force participation rate $p_{ai}$ for the relevant age group using the latest available country-specific data (indicator *EAP_DWAP_SEX_AGE_RT_A* from the ILOSTAT library).^11^ These data are available in five-year age intervals: 15–19, 20–24, 25–29, 30–34, 35–39, 40–44, 45–49, 50–54, 55–59, 60–64, ≥65. The size of the labor force in each age group is therefore calculated as $L_{ait}=p_{ai}N_{ait}$.

These age-specific stocks of labor were then augmented by experience using a standard Cuddington and Hancock (1994) specification.^13^ Skill-augmented labor $L_{it}$ is defined as:

$$L_{it}=\sum_{a=15}^{60} \rho_{a}L_{ait},$$

(3)

where $\rho_{a}$ is the Cuddington/skill-augmenting factor that adjusts for skill and experience for different ages and was defined as:

$$\rho_{a}=\rho_{1}+\rho_{2}\left( {age}_{a}-15 \right)+\rho_{3}({age}_{a}-15)^{2},$$

(4)

where ${age}_{a}$ is the age of a worker while $\rho_{1}$ = 0.8, $\rho_{2}$ = 0.02, and $\rho_{3}$ = -0.0002.

Educational capital and total factor productivity were assumed to grow at rates $h_{it}$ and $g_{it}$, respectively:

$$H_{it}=H_{i(t-1)}e^{h_{it}}, A_{it}=A_{i(t-1)}e^{g_{it}}.$$

(5)

The growth rates of educational attainment and total factor productivity were computed from the levels reported in the Penn World Tables.^10,14,15^ The specific PWT variables from which we computed the annual growth rates were *rtfpna* (for $g_{it}$) and *hc* (for $h_{it}$). The educational attainment index *hc* was based on years of schooling^14^ and returns to education.^15^ The construction of total factor productivity *rtfpna* in the PWT is detailed in.^10^

When costs are introduced, the evolution of physical capital can be rewritten as:

$K_{it}=s_{it}Y_{i(t-1)}-C_{it}+(1-\delta_{it})K_{i(t-1)}$ ,

(6)

where $C_{it}$ is the public sector cost of TB vaccination (net of international donor contributions), net of patient out-of-pocket and public expenditure treatment savings.

Improved health status stemming from the intervention has positive effects on the size of the effective labor force due to reduced mortality, as established by the *Mtb* transmission model. This counterfactual labor force is constructed by replacing the projected $N_{ait}$ series estimated by the transmission model with the simulation projection $\tilde{N}_{ait}$ and proceeding as detailed above.

The increase in the projected labor force by $\tilde{L}_{it}-L_{it}$ is then the main source of gains in GDP from the novel TB vaccine intervention, which is mitigated when net intervention costs $C_{it}$ are strictly positive.

The initial level of the stock of physical capital per efficiency unit of labor is calibrated so as to correspond to its steady-state level $k_{it}^{*}$ given by:

$$k_{it}^{*}=(\frac{s_{it}\gamma_{i}A_{it}}{n_{it}+h_{it}+\delta_{it}})^{\frac{1}{1-\alpha_{it}}} ,$$

(7)

where $k\equiv K/HL$. This avoids unrealistically high growth rates generated by what would correspond to transitional dynamics toward the steady-state at the start of the simulation, and therefore yields the balanced growth path.

### Appendix S3. Cumulative absolute gains to gross domestic product (GDP, billions US$2020) due to adolescent/adult tuberculosis vaccines by decade.

| Country grouping | 2041–2050 | 2051–2060 | 2061–2070 | 2071–2080 |
| --- | --- | --- | --- | --- |
| All countries | 111 (43.5, 201) | 283 (126, 528) | 482 (204, 955) | 729 (284, 1518) |
| High-TB burden^a^ | 113 (54.0, 197) | 280 (135, 511) | 471 (212, 920) | 708 (290, 1457) |
| High-TB/HIV burden^a^ | 105 (54.9, 176) | 254 (127, 453) | 420 (194, 804) | 621 (262, 1250) |
| High-MDR/RR-TB burden^a^ | 109 (52.6, 190) | 269 (130, 491) | 451 (203, 883) | 678 (277, 1397) |
|  | Income level^b^ | | | |
| LIC | 2.58 (0.61, 5.19) | 8.30 (2.88, 16.3) | 15.9 (5.32, 32.7) | 26.5 (8.24, 56.7) |
| LMIC | 97.2 (49.6, 165) | 248 (122, 449) | 423 (191, 824) | 642 (262, 1320) |
| UMIC | 11.2 (-6.70, 30.2) | 27.0 (1.34, 62.2) | 42.5 (7.26, 97.9) | 60.0 (13.2, 141) |
|  | World region | | | |
| AFR | 36.7 (19.8, 60.6) | 77.6 (40.0, 135) | 114 (54.1, 212) | 151 (66.1, 296) |
| AMR | -2.07 (-5.34, -0.37) | -1.54 (-4.83, 0.44) | -1.16 (-4.43, 1.07) | -0.90 (-4.17, 1.51) |
| EMR | -1.17 (-3.65, 0.96) | 1.33 (-2.31, 5.32) | 3.39 (-1.56, 9.51) | 5.27 (-1.02, 13.5) |
| EUR | -0.64 (-2.97, 0.96) | 0.11 (-2.86, 2.80) | 0.86 (-3.06, 5.13) | 1.95 (-3.19, 8.53) |
| SEAR | 64.7 (33.9, 110) | 169 (83.2, 308) | 293 (133, 573) | 448 (184, 915) |
| WPR | 13.4 (1.74, 29.1) | 36.8 (12.9, 76.3) | 71.4 (26.0, 155) | 123 (42.3, 284) |

Note: Values in parentheses represent equal-tailed 95% uncertainty intervals.

^a^ High-TB, high-TB/HIV (HIV-associated TB), and high-MDR/RR-TB (multidrug/rifampicin-resistant TB) burden countries as defined by the World Health Organization.

^b^ LIC: Gross national income (GNI) per capita of $1,085 or less; LMIC: GNI per capita of $1,086 to $4,225; UMIC: GNI per capita of $4,256 to $13,205 (World Bank 2021).

Note: All countries include 105 low- and middle-income countries analyzed. AFR = African region; AMR = Region of the Americas; EMR = Eastern Mediterranean region; EUR = European region; GDP = gross domestic product; LIC = low-income; LMIC = lower middle-income; SEAR = Southeast Asian region; UMIC = upper middle-income; WPR = Western Pacific region.

### Appendix S4. Cumulative absolute gains to gross domestic product (GDP, billions US$2020) due to infant tuberculosis vaccines by decade.

| Country grouping | 2041–2050 | 2051–2060 | 2061–2070 | 2071–2080 |
| --- | --- | --- | --- | --- |
| All countries | -0.75 (-5.17, 3.62) | 18.4 (0.75, 44.1) | 62.0 (16.1, 138) | 128 (36.1, 288) |
| High-TB burden^a^ | 0.98 (-2.10, 4.60) | 21.6 (6.97, 45.3) | 65.9 (25, 138) | 132 (47.6, 286) |
| High-TB/HIV burden^a^ | 1.51 (-0.86, 4.59) | 19.3 (6.86, 39.6) | 57.3 (22.4, 118) | 112 (41.7, 239) |
| High-MDR/RR-TB burden^a^ | 1.10 (-1.88, 4.62) | 20.9 (6.71, 43.9) | 63.5 (23.9, 133) | 127 (45.6, 275) |
|  | Income level^b^ | | | |
| LIC | -0.26 (-0.46, -0.09) | -0.003 (-0.80, 0.93) | 0.96 (-0.96, 3.59) | 2.79 (-1.03, 8.47) |
| LMIC | 0.97 (-1.47, 4.06) | 17.9 (5.01, 38.3) | 55.6 (19.5, 119) | 113 (38.4, 247) |
| UMIC | -1.46 (-3.24, -0.35) | 0.55 (-3.46, 4.82) | 5.43 (-2.47, 16.0) | 12.2 (-1.21, 32.7) |
|  | World region | | | |
| AFR | 0.66 (-0.67, 2.42) | 7.67 (1.77, 16.9) | 19.4 (6.11, 41.6) | 32.7 (10.0, 70.5) |
| AMR | -0.52 (-0.92, -0.30) | -0.83 (-1.56, -0.44) | -1.02 (-2.01, -0.50) | -1.16 (-2.35, -0.51) |
| EMR | -0.67 (-1.11, -0.37) | -0.71 (-1.77, 0.18) | -0.50 (-2.22, 1.17) | -0.17 (-2.62, 2.39) |
| EUR | -0.41 (-0.80, -0.22) | -0.70 (-1.46, -0.32) | -0.84 (-1.97, -0.22) | -0.88 (-2.43, 0.12) |
| SEAR | 0.52 (-0.38, 1.66) | 9.27 (3.40, 19.0) | 31.3 (12.3, 65.1) | 66.2 (24.5, 142) |
| WPR | -0.33 (-1.28, 0.42) | 3.70 (0.36, 8.73) | 13.7 (3.90, 30.9) | 31.0 (9.05, 73.8) |

Note: Values in parentheses represent equal-tailed 95% uncertainty intervals.

^a^ High-TB, high-TB/HIV (HIV-associated TB), and high-MDR/RR-TB (multidrug/rifampicin-resistant TB) burden countries as defined by the World Health Organization.

^b^ LIC: Gross national income (GNI) per capita of $1,085 or less; LMIC: GNI per capita of $1,086 to $4,225; UMIC: GNI per capita of $4,256 to $13,205 (World Bank 2021).

Note: All countries include 105 low- and middle-income countries analyzed. AFR = African region; AMR = Region of the Americas; EMR = Eastern Mediterranean region; EUR = European region; GDP = gross domestic product; LIC = low-income; LMIC = lower middle-income; SEAR = Southeast Asian region; UMIC = upper middle-income; WPR = Western Pacific region.

### Appendix S5. Gains to gross domestic product (GDP) due to adolescent/adult tuberculosis vaccines across 2028–2080: health services costs included government-level and excluded patient- and international-donor-level.

| Country grouping | Absolute gains in GDP  (billions US$2020) | Percentage gain in GDP  (%) |
| --- | --- | --- |
| All countries | 1552 (715, 2900) | 0.0311% (0.0254%, 0.0372%) |
| High-TB burden^a^ | 1528 (716, 2833) | 0.0380% (0.0316%, 0.0452%) |
| High-TB/HIV burden^a^ | 1366 (653, 2498) | 0.0823% (0.0693%, 0.0974%) |
| High-MDR/RR-TB burden^a^ | 1469 (690, 2710) | 0.0368% (0.0305%, 0.0439%) |
|  | Income level^b^ | |
| LIC | 43.0 (16.3, 90.7) | 0.0308% (0.0213%, 0.0400%) |
| LMIC | 1371 (637, 2531) | 0.0810% (0.0690%, 0.0955%) |
| UMIC | 138 (35.5, 294) | 0.0043% (0.0021%, 0.0058%) |
|  | World region | |
| AFR | 367 (189, 654) | 0.1105% (0.0942%, 0.1279%) |
| AMR | -6.29 (-18.2, -0.45) | -0.0021% (-0.0058%, -0.0001%) |
| EMR | -1.37 (-9.74, 8.59) | -0.0016% (-0.0103%, 0.0046%) |
| EUR | 0.95 (-9.28, 11.4) | 0.00004% (-0.0028%, 0.0015%) |
| SEAR | 957 (439, 1785) | 0.0823% (0.0667%, 0.1003%) |
| WPR | 235 (86.6, 495) | 0.0089% (0.0064%, 0.0115%) |

Note: Values in parentheses represent equal-tailed 95% uncertainty intervals.

^a^ High-TB, high-TB/HIV (HIV-associated TB), and high-MDR/RR-TB (multidrug/rifampicin-resistant TB) burden countries as defined by the World Health Organization.

^b^ LIC: Gross national income (GNI) per capita of $1,085 or less; LMIC: GNI per capita of $1,086 to $4,225; UMIC: GNI per capita of $4,256 to $13,205 (World Bank 2021).

Note: All countries include 105 low- and middle-income countries analyzed. AFR = African region; AMR = Region of the Americas; EMR = Eastern Mediterranean region; EUR = European region; GDP = gross domestic product; LIC = low-income; LMIC = lower middle-income; SEAR = Southeast Asian region; UMIC = upper middle-income; WPR = Western Pacific region.

### Appendix S6. Gains to gross domestic product (GDP) due to infant tuberculosis vaccines across 2028–2080: health services costs included government-level and excluded patient- and international-donor-level.

| Country grouping | Absolute gains in GDP  (billions US$2020) | Percentage gain in GDP  (%) |
| --- | --- | --- |
| All countries | 198 (73.0, 395) | 0.0039% (0.0026%, 0.0052%) |
| High-TB burden^a^ | 211 (85.1, 406) | 0.0052% (0.0038%, 0.0066%) |
| High-TB/HIV burden^a^ | 184 (78.3, 345) | 0.0110% (0.0079%, 0.0143%) |
| High-MDR/RR-TB burden^a^ | 204 (82.7, 393) | 0.0051% (0.0037%, 0.0064%) |
| Income level^b^ | Income level^b^ | |
| LIC | 1.98 (-1.37, 8.05) | 0.0011% (-0.0017%, 0.0035%) |
| LMIC | 180 (72.7, 349) | 0.0105% (0.0075%, 0.0138%) |
| UMIC | 16.4 (-0.94, 43.7) | 0.0005% (-0.00004%, 0.0008%) |
| World region | World region | |
| AFR | 58.4 (24.1, 116) | 0.0174% (0.0114%, 0.0243%) |
| AMR | -3.35 (-5.82, -2.17) | -0.0011% (-0.0019%, -0.0006%) |
| EMR | -4.35 (-7.05, -2.21) | -0.0034% (-0.0064%, -0.0012%) |
| EUR | -2.74 (-5.5, -1.46) | -0.0007% (-0.0016%, -0.0003%) |
| SEAR | 104 (42.2, 207) | 0.0089% (0.0060%, 0.0125%) |
| WPR | 46.0 (14.6, 102) | 0.0017% (0.0010%, 0.0025%) |

Note: Values in parentheses represent equal-tailed 95% uncertainty intervals.

^a^ High-TB, high-TB/HIV (HIV-associated TB), and high-MDR/RR-TB (multidrug/rifampicin-resistant TB) burden countries as defined by the World Health Organization.

^b^ LIC: Gross national income (GNI) per capita of $1,085 or less; LMIC: GNI per capita of $1,086 to $4,225; UMIC: GNI per capita of $4,256 to $13,205 (World Bank 2021).

Note: All countries include 105 low- and middle-income countries analyzed. AFR = African region; AMR = Region of the Americas; EMR = Eastern Mediterranean region; EUR = European region; GDP = gross domestic product; LIC = low-income; LMIC = lower middle-income; SEAR = Southeast Asian region; UMIC = upper middle-income; WPR = Western Pacific region.

### Appendix S7. Gains to gross domestic product (GDP) due to adolescent/adult tuberculosis vaccines across 2028–2080: health services costs included patient-level and excluded government- and international-donor-level.

| Country grouping | Absolute gains in GDP  (billions US$2020) | Percentage gain in GDP  (%) |
| --- | --- | --- |
| All countries | 1772 (875, 3211) | 0.0358% (0.0305%, 0.0421%) |
| High-TB burden^a^ | 1685 (835, 3049) | 0.0421% (0.0358%, 0.0494%) |
| High-TB/HIV burden^a^ | 1470 (741, 2649) | 0.0890% (0.0765%, 0.1035%) |
| High-MDR/RR-TB burden^a^ | 1615 (799, 2911) | 0.0407% (0.0345%, 0.0478%) |
|  | Income level^b^ | |
| LIC | 67.9 (35.3, 123) | 0.0503% (0.0435%, 0.0577%) |
| LMIC | 1498 (741, 2713) | 0.0889% (0.0771%, 0.1030%) |
| UMIC | 206 (97.1, 374) | 0.0066% (0.0056%, 0.0076%) |
|  | World region | |
| AFR | 418 (228, 728) | 0.1268% (0.1110%, 0.1441%) |
| AMR | 8.67 (4.39, 15.0) | 0.0027% (0.0021%, 0.0032%) |
| EMR | 26.9 (15.6, 44.1) | 0.0197% (0.0149%, 0.0256%) |
| EUR | 15.0 (7.38, 27.1) | 0.0035% (0.0029%, 0.0041%) |
| SEAR | 1015 (484, 1862) | 0.0876% (0.0719%, 0.1052%) |
| WPR | 288 (127, 551) | 0.0111% (0.0092%, 0.0133%) |

Note: Values in parentheses represent equal-tailed 95% uncertainty intervals.

^a^ High-TB, high-TB/HIV (HIV-associated TB), and high-MDR/RR-TB (multidrug/rifampicin-resistant TB) burden countries as defined by the World Health Organization.

^b^ LIC: Gross national income (GNI) per capita of $1,085 or less; LMIC: GNI per capita of $1,086 to $4,225; UMIC: GNI per capita of $4,256 to $13,205 (World Bank 2021).

Note: All countries include 105 low- and middle-income countries analyzed. AFR = African region; AMR = Region of the Americas; EMR = Eastern Mediterranean region; EUR = European region; GDP = gross domestic product; LIC = low-income; LMIC = lower middle-income; SEAR = Southeast Asian region; UMIC = upper middle-income; WPR = Western Pacific region.

### Appendix S8. Gains to gross domestic product (GDP) due to infant tuberculosis vaccines across 2028–2080: health services costs included patient-level and excluded government- and international-donor-level.

| Country grouping | Absolute gains in GDP  (billions US$2020) | Percentage gain in GDP  (%) |
| --- | --- | --- |
| All countries | 257 (120, 470) | 0.0052% (0.0041%, 0.0064%) |
| High-TB burden^a^ | 249 (116, 456) | 0.0062% (0.0050%, 0.0077%) |
| High-TB/HIV burden^a^ | 211 (99.8, 380) | 0.0127% (0.0100%, 0.0160%) |
| High-MDR/RR-TB burden^a^ | 239 (111, 438) | 0.0060% (0.0048%, 0.0075%) |
| Income level^b^ | Income level^b^ | |
| LIC | 9.39 (4.74, 17.1) | 0.0069% (0.0057%, 0.0083%) |
| LMIC | 214 (99.5, 395) | 0.0127% (0.0101%, 0.0158%) |
| UMIC | 33.1 (14.2, 63.5) | 0.0010% (0.0008%, 0.0013%) |
| World region | World region | |
| AFR | 76.5 (38.6, 140) | 0.0231% (0.0178%, 0.0299%) |
| AMR | 0.53 (0.17, 1.06) | 0.0002% (0.0001%, 0.0002%) |
| EMR | 5.33 (2.94, 8.98) | 0.0039% (0.0027%, 0.0056%) |
| EUR | 1.07 (0.38, 2.35) | 0.0002% (0.0002%, 0.0003%) |
| SEAR | 116 (51.9, 220) | 0.0100% (0.0073%, 0.0135%) |
| WPR | 57.3 (23.6, 116) | 0.0022% (0.0016%, 0.0029%) |

Note: Values in parentheses represent equal-tailed 95% uncertainty intervals.

^a^ High-TB, high-TB/HIV (HIV-associated TB), and high-MDR/RR-TB (multidrug/rifampicin-resistant TB) burden countries as defined by the World Health Organization.

^b^ LIC: Gross national income (GNI) per capita of $1,085 or less; LMIC: GNI per capita of $1,086 to $4,225; UMIC: GNI per capita of $4,256 to $13,205 (World Bank 2021).

Note: All countries include 105 low- and middle-income countries analyzed. AFR = African region; AMR = Region of the Americas; EMR = Eastern Mediterranean region; EUR = European region; GDP = gross domestic product; LIC = low-income; LMIC = lower middle-income; SEAR = Southeast Asian region; UMIC = upper middle-income; WPR = Western Pacific region.

### Appendix S9. Gains to gross domestic product (GDP) due to adolescent/adult tuberculosis vaccines across 2028–2080: health services costs included all levels (government, patient, international donor).

| Country grouping | Absolute gains in GDP  (billions US$2020) | Percentage gain in GDP  (%) |
| --- | --- | --- |
| All countries | 1523 (698, 2849) | 0.0306% (0.0247%, 0.0367%) |
| High-TB burden^a^ | 1536 (720, 2833) | 0.0382% (0.0314%, 0.0455%) |
| High-TB/HIV burden^a^ | 1368 (654, 2491) | 0.0825% (0.0694%, 0.0977%) |
| High-MDR/RR-TB burden^a^ | 1483 (698, 2715) | 0.0372% (0.0305%, 0.0445%) |
|  | Income level^b^ | |
| LIC | 11.8 (-16.1, 50.1) | 0.0067% (-0.0188%, 0.0227%) |
| LMIC | 1369 (642, 2516) | 0.0809% (0.0681%, 0.0954%) |
| UMIC | 142.9 (38.6, 303) | 0.0044% (0.0022%, 0.0060%) |
|  | World region | |
| AFR | 337 (167, 607) | 0.1011% (0.0801%, 0.1200%) |
| AMR | -7.20 (-20.4, -1.10) | -0.0024% (-0.0065%, -0.0002%) |
| EMR | -7.30 (-21.7, 5.22) | -0.0060% (-0.0194%, 0.0028%) |
| EUR | -0.25 (-11.6, 10.1) | -0.00025% (-0.0033%, 0.0014%) |
| SEAR | 958 (441, 1781) | 0.0824% (0.0665%, 0.1009%) |
| WPR | 244 (92.3, 504) | 0.0093% (0.0066%, 0.0118%) |

Note: Values in parentheses represent equal-tailed 95% uncertainty intervals.

^a^ High-TB, high-TB/HIV (HIV-associated TB), and high-MDR/RR-TB (multidrug/rifampicin-resistant TB) burden countries as defined by the World Health Organization.

^b^ LIC: Gross national income (GNI) per capita of $1,085 or less; LMIC: GNI per capita of $1,086 to $4,225; UMIC: GNI per capita of $4,256 to $13,205 (World Bank 2021).

Note: All countries include 105 low- and middle-income countries analyzed. AFR = African region; AMR = Region of the Americas; EMR = Eastern Mediterranean region; EUR = European region; GDP = gross domestic product; LIC = low-income; LMIC = lower middle-income; SEAR = Southeast Asian region; UMIC = upper middle-income; WPR = Western Pacific region.

### Appendix S10. Gains to gross domestic product (GDP) due to infant tuberculosis vaccines across 2028–2080: health services costs included all levels (government, patient, international donor).

| Country grouping | Absolute gains in GDP  (billions US$2020) | Percentage gain in GDP  (%) |
| --- | --- | --- |
| All countries | 171 (51.1, 370) | 0.0033% (0.0018%, 0.0047%) |
| High-TB burden^a^ | 201 (77.4, 391) | 0.0049% (0.0034%, 0.0064%) |
| High-TB/HIV burden^a^ | 173 (69.2, 334) | 0.0103% (0.0071%, 0.0137%) |
| High-MDR/RR-TB burden^a^ | 199 (78.9, 387) | 0.0049% (0.0035%, 0.0064%) |
| Income level^b^ | Income level^b^ | |
| LIC | -13.1 (-20.4, -8.30) | -0.0106% (-0.0201%, -0.0043%) |
| LMIC | 168 (64.3, 335) | 0.0098% (0.0065%, 0.0132%) |
| UMIC | 16.3 (-1.28, 43.7) | 0.0005% (-0.0001%, 0.0008%) |
| World region | World region | |
| AFR | 55.8 (20.7, 111) | 0.0168% (0.0096%, 0.0242%) |
| AMR | -3.58 (-6.33, -2.27) | -0.0012% (-0.0021%, -0.0007%) |
| EMR | -5.96 (-11.2, -2.23) | -0.0046% (-0.0092%, -0.0014%) |
| EUR | -3.21 (-6.37, -1.74) | -0.0008% (-0.0018%, -0.0003%) |
| SEAR | 105 (42.5, 208) | 0.0089% (0.0060%, 0.0126%) |
| WPR | 49.7 (17.0, 107) | 0.0019% (0.0012%, 0.0026%) |

Note: Values in parentheses represent equal-tailed 95% uncertainty intervals.

^a^ High-TB, high-TB/HIV (HIV-associated TB), and high-MDR/RR-TB (multidrug/rifampicin-resistant TB) burden countries as defined by the World Health Organization.

^b^ LIC: Gross national income (GNI) per capita of $1,085 or less; LMIC: GNI per capita of $1,086 to $4,225; UMIC: GNI per capita of $4,256 to $13,205 (World Bank 2021).

Note: All countries include 105 low- and middle-income countries analyzed. AFR = African region; AMR = Region of the Americas; EMR = Eastern Mediterranean region; EUR = European region; GDP = gross domestic product; LIC = low-income; LMIC = lower middle-income; SEAR = Southeast Asian region; UMIC = upper middle-income; WPR = Western Pacific region.

### Appendix S11. Gains to gross domestic product (GDP) due to adolescent/adult tuberculosis vaccines across 2028–2080: low-growth scenario.

| Country grouping | Absolute gains in GDP  (billions US$2020) | Percentage gain in GDP  (%) |
| --- | --- | --- |
| All countries | 1060 (812, 1399) | 0.0312% (0.0255%, 0.0385%) |
| High-TB burden^a^ | 1049 (802, 1386) | 0.0383% (0.0304%, 0.0494%) |
| High-TB/HIV burden^a^ | 948 (727, 1245) | 0.0816% (0.0688%, 0.0962%) |
| High-MDR/RR-TB burden^a^ | 1009 (766, 1342) | 0.0372% (0.0293%, 0.0475%) |
|  | Income level^b^ | |
| LIC | 32.6 (24.2, 41.0) | 0.0371% (0.0307%, 0.0443%) |
| LMIC | 941 (698, 1263) | 0.0815% (0.0690%, 0.0954%) |
| UMIC | 87.0 (44.8, 138) | 0.0040% (0.0022%, 0.0054%) |
|  | World region | |
| AFR | 269 (194, 356) | 0.1171% (0.0975%, 0.1419%) |
| AMR | -6.40 (-12.4, -3.20) | -0.0026% (-0.0053%, -0.0012%) |
| EMR | 4.00 (-2.13, 10.4) | 0.0041% (-0.0024%, 0.0107%) |
| EUR | -0.50 (-5.79, 3.21) | -0.0002% (-0.0021%, 0.0011%) |
| SEAR | 648 (413, 994) | 0.0810% (0.0645%, 0.0996%) |
| WPR | 146 (89.5, 203) | 0.0085% (0.0061%, 0.0120%) |

Note: Values in parentheses represent equal-tailed 95% uncertainty intervals.

^a^ High-TB, high-TB/HIV (HIV-associated TB), and high-MDR/RR-TB (multidrug/rifampicin-resistant TB) burden countries as defined by the World Health Organization.

^b^ LIC: Gross national income (GNI) per capita of $1,085 or less; LMIC: GNI per capita of $1,086 to $4,225; UMIC: GNI per capita of $4,256 to $13,205 (World Bank 2021).

Note: All countries include 105 low- and middle-income countries analyzed. AFR = African region; AMR = Region of the Americas; EMR = Eastern Mediterranean region; EUR = European region; GDP = gross domestic product; LIC = low-income; LMIC = lower middle-income; SEAR = Southeast Asian region; UMIC = upper middle-income; WPR = Western Pacific region.

### Appendix S12. Gains to gross domestic product (GDP) due to infant tuberculosis vaccines across 2028–2080: low-growth scenario.

| Country grouping | Absolute gains in GDP  (billions US$2020) | Percentage gain in GDP  (%) |
| --- | --- | --- |
| All countries | 125 (82.6, 178) | 0.0037% (0.0026%, 0.0049%) |
| High-TB burden^a^ | 137 (95.4, 188) | 0.0050% (0.0037%, 0.0068%) |
| High-TB/HIV burden^a^ | 122 (82.8, 170) | 0.0105% (0.0080%, 0.0135%) |
| High-MDR/RR-TB burden^a^ | 133 (90.8, 184) | 0.0049% (0.0036%, 0.0067%) |
| Income level^b^ | Income level^b^ | |
| LIC | 1.19 (-0.18, 2.62) | 0.0013% (-0.0002%, 0.0027%) |
| LMIC | 118 (78.4, 167) | 0.0102% (0.0077%, 0.0132%) |
| UMIC | 6.72 (-1.74, 16.2) | 0.0003% (-0.0001%, 0.0006%) |
| World region | World region | |
| AFR | 40.1 (20.8, 66.1) | 0.0174% (0.0103%, 0.0262%) |
| AMR | -3.10 (-4.30, -2.44) | -0.0013% (-0.0018%, -0.0009%) |
| EMR | -2.30 (-4.21, -0.49) | -0.0024% (-0.0044%, -0.0005%) |
| EUR | -2.63 (-4.04, -1.78) | -0.0009% (-0.0016%, -0.0005%) |
| SEAR | 67.6 (37.9, 109) | 0.0084% (0.0058%, 0.0123%) |
| WPR | 25.7 (13.9, 38.6) | 0.0015% (0.0009%, 0.0021%) |

Note: Values in parentheses represent equal-tailed 95% uncertainty intervals.

^a^ High-TB, high-TB/HIV (HIV-associated TB), and high-MDR/RR-TB (multidrug/rifampicin-resistant TB) burden countries as defined by the World Health Organization.

^b^ LIC: Gross national income (GNI) per capita of $1,085 or less; LMIC: GNI per capita of $1,086 to $4,225; UMIC: GNI per capita of $4,256 to $13,205 (World Bank 2021).

Note: All countries include 105 low- and middle-income countries analyzed. AFR = African region; AMR = Region of the Americas; EMR = Eastern Mediterranean region; EUR = European region; GDP = gross domestic product; LIC = low-income; LMIC = lower middle-income; SEAR = Southeast Asian region; UMIC = upper middle-income; WPR = Western Pacific region.

### Appendix S13. Gains to gross domestic product (GDP) due to adolescent/adult tuberculosis vaccines across 2028–2080: high-growth scenario.

| Country grouping | Absolute gains in GDP  (billions US$2020) | Percentage gain in GDP  (%) |
| --- | --- | --- |
| All countries | 2295 (1796, 3036) | 0.0345% (0.0249%, 0.0457%) |
| High-TB burden^a^ | 2239 (1740, 2974) | 0.0418% (0.0283%, 0.0560%) |
| High-TB/HIV burden^a^ | 1972 (1489, 2712) | 0.0897% (0.0779%, 0.1064%) |
| High-MDR/RR-TB burden^a^ | 2142 (1653, 2831) | 0.0404% (0.0273%, 0.0542%) |
|  | Income level^b^ | |
| LIC | 80.3 (59.9, 114) | 0.0412% (0.0338%, 0.0493%) |
| LMIC | 2004 (1523, 2686) | 0.0872% (0.0750%, 0.1021%) |
| UMIC | 211 (128, 371) | 0.0050% (0.0038%, 0.0063%) |
|  | World region | |
| AFR | 525 (404, 715) | 0.1163% (0.0991%, 0.1406%) |
| AMR | -6.46 (-13.8, -1.49) | -0.0016% (-0.0036%, -0.0003%) |
| EMR | 13.8 (3.23, 27.2) | 0.0074% (0.0017%, 0.0138%) |
| EUR | 4.81 (-3.20, 15.7) | 0.0008% (-0.0006%, 0.0022%) |
| SEAR | 1383 (978, 1947) | 0.0884% (0.0721%, 0.1077%) |
| WPR | 375 (259, 593) | 0.0108% (0.0079%, 0.0156%) |

Note: Values in parentheses represent equal-tailed 95% uncertainty intervals.

^a^ High-TB, high-TB/HIV (HIV-associated TB), and high-MDR/RR-TB (multidrug/rifampicin-resistant TB) burden countries as defined by the World Health Organization.

^b^ LIC: Gross national income (GNI) per capita of $1,085 or less; LMIC: GNI per capita of $1,086 to $4,225; UMIC: GNI per capita of $4,256 to $13,205 (World Bank 2021).

Note: All countries include 105 low- and middle-income countries analyzed. AFR = African region; AMR = Region of the Americas; EMR = Eastern Mediterranean region; EUR = European region; GDP = gross domestic product; LIC = low-income; LMIC = lower middle-income; SEAR = Southeast Asian region; UMIC = upper middle-income; WPR = Western Pacific region.

### Appendix S14. Gains to gross domestic product (GDP) due to infant tuberculosis vaccines across 2028–2080: high-growth scenario.

| Country grouping | Absolute gains in GDP  (billions US$2020) | Percentage gain in GDP  (%) |
| --- | --- | --- |
| All countries | 306 (223, 440) | 0.0046% (0.0032%, 0.0063%) |
| High-TB burden^a^ | 320 (239, 455) | 0.0060% (0.0040%, 0.0085%) |
| High-TB/HIV burden^a^ | 273 (189, 407) | 0.0124% (0.0099%, 0.0162%) |
| High-MDR/RR-TB burden^a^ | 308 (225, 434) | 0.0058% (0.0038%, 0.0084%) |
| Income level^b^ | Income level^b^ | |
| LIC | 6.55 (2.88, 11.9) | 0.0033% (0.0017%, 0.0050%) |
| LMIC | 272 (187, 395) | 0.0119% (0.0092%, 0.0156%) |
| UMIC | 27.6 (12.2, 57.5) | 0.0006% (0.0004%, 0.0009%) |
| World region | World region | |
| AFR | 85.2 (55.9, 143) | 0.0188% (0.0132%, 0.0285%) |
| AMR | -4.32 (-6.10, -3.17) | -0.0011% (-0.0016%, -0.0007%) |
| EMR | -1.87 (-5.42, 1.39) | -0.001% (-0.0029%, 0.0008%) |
| EUR | -3.36 (-6.35, -1.73) | -0.0006% (-0.0013%, -0.0003%) |
| SEAR | 155 (96.2, 231) | 0.0099% (0.0070%, 0.0138%) |
| WPR | 75.6 (45.9, 128) | 0.0022% (0.0015%, 0.0031%) |

Note: Values in parentheses represent equal-tailed 95% uncertainty intervals.

^a^ High-TB, high-TB/HIV (HIV-associated TB), and high-MDR/RR-TB (multidrug/rifampicin-resistant TB) burden countries as defined by the World Health Organization.

^b^ LIC: Gross national income (GNI) per capita of $1,085 or less; LMIC: GNI per capita of $1,086 to $4,225; UMIC: GNI per capita of $4,256 to $13,205 (World Bank 2021).

Note: All countries include 105 low- and middle-income countries analyzed. AFR = African region; AMR = Region of the Americas; EMR = Eastern Mediterranean region; EUR = European region; GDP = gross domestic product; LIC = low-income; LMIC = lower middle-income; SEAR = Southeast Asian region; UMIC = upper middle-income; WPR = Western Pacific region.

### Appendix S15. Gains to gross domestic product (GDP) due to adolescent/adult tuberculosis vaccines across 2028–2080: scenario with alternative approach to modelling the consequences of TB morbidity outcomes.

| Country grouping | Absolute gains in GDP  (billions US$2020) | Percentage gain in GDP  (%) |
| --- | --- | --- |
| All countries | 1338 (654, 2422) | 0.0270% (0.0221%, 0.0322%) |
| High-TB burden^a^ | 1316 (654, 2364) | 0.0329% (0.0273%, 0.0390%) |
| High-TB/HIV burden^a^ | 1199 (606, 2113) | 0.0726% (0.0618%, 0.0839%) |
| High-MDR/RR-TB burden^a^ | 1256 (623, 2247) | 0.0317% (0.0262%, 0.0377%) |
|  | Income level^b^ | |
| LIC | 51.0 (26.2, 89.6) | 0.0378% (0.0306%, 0.0451%) |
| LMIC | 1183 (583, 2111) | 0.0702% (0.0602%, 0.0809%) |
| UMIC | 103 (20.9, 222) | 0.0032% (0.0009%, 0.0045%) |
|  | World region | |
| AFR | 345 (189, 590) | 0.1047% (0.0901%, 0.1197%) |
| AMR | -7.43 (-20.2, -1.78) | -0.0024% (-0.0064%, -0.0005%) |
| EMR | 9.91 (-0.01, 21.8) | 0.0071% (-0.00001%, 0.013%) |
| EUR | -1.21 (-11.4, 6.28) | -0.0004% (-0.0031%, 0.0010%) |
| SEAR | 813 (387, 1474) | 0.0701% (0.0571%, 0.0837%) |
| WPR | 178 (70.7, 364) | 0.0068% (0.0049%, 0.0086%) |

Note: Values in parentheses represent equal-tailed 95% uncertainty intervals.

^a^ High-TB, high-TB/HIV (HIV-associated TB), and high-MDR/RR-TB (multidrug/rifampicin-resistant TB) burden countries as defined by the World Health Organization.

^b^ LIC: Gross national income (GNI) per capita of $1,085 or less; LMIC: GNI per capita of $1,086 to $4,225; UMIC: GNI per capita of $4,256 to $13,205 (World Bank 2021).

Note: All countries include 105 low- and middle-income countries analyzed. AFR = African region; AMR = Region of the Americas; EMR = Eastern Mediterranean region; EUR = European region; GDP = gross domestic product; LIC = low-income; LMIC = lower middle-income; SEAR = Southeast Asian region; UMIC = upper middle-income; WPR = Western Pacific region.

### Appendix S16. Gains to gross domestic product (GDP) due to infant tuberculosis vaccines across 2028–2080: scenario with alternative approach to modelling the consequences of TB morbidity outcomes.

| Country grouping | Absolute gains in GDP  (billions US$2020) | Percentage gain in GDP  (%) |
| --- | --- | --- |
| All countries | 187 (73.6, 364) | 0.0037% (0.0026%, 0.0048%) |
| High-TB burden^a^ | 198 (84.8, 370) | 0.0049% (0.0037%, 0.0062%) |
| High-TB/HIV burden^a^ | 173 (76.7, 316) | 0.0104% (0.0078%, 0.0131%) |
| High-MDR/RR-TB burden^a^ | 191 (81.8, 353) | 0.0048% (0.0035%, 0.006%) |
| Income level^b^ | Income level^b^ | |
| LIC | 4.38 (0.82, 10.2) | 0.0031% (0.0009%, 0.0047%) |
| LMIC | 168 (72.1, 316) | 0.0099% (0.0073%, 0.0126%) |
| UMIC | 14.5 (-2.00, 38.1) | 0.0004% (-0.0001%, 0.0008%) |
| World region | World region | |
| AFR | 55.1 (23.9, 107) | 0.0165% (0.0114%, 0.0227%) |
| AMR | -3.53 (-6.22, -2.25) | -0.0011% (-0.0021%, -0.0007%) |
| EMR | -1.08 (-4.10, 1.58) | -0.0009% (-0.0035%, 0.0011%) |
| EUR | -2.97 (-6.02, -1.58) | -0.0008% (-0.0017%, -0.0003%) |
| SEAR | 96.0 (40.0, 186) | 0.0082% (0.0057%, 0.0114%) |
| WPR | 43.0 (14.6, 91.9) | 0.0016% (0.0010%, 0.0023%) |

Note: Values in parentheses represent equal-tailed 95% uncertainty intervals.

^a^ High-TB, high-TB/HIV (HIV-associated TB), and high-MDR/RR-TB (multidrug/rifampicin-resistant TB) burden countries as defined by the World Health Organization.

^b^ LIC: Gross national income (GNI) per capita of $1,085 or less; LMIC: GNI per capita of $1,086 to $4,225; UMIC: GNI per capita of $4,256 to $13,205 (World Bank 2021).

Note: All countries include 105 low- and middle-income countries analyzed. AFR = African region; AMR = Region of the Americas; EMR = Eastern Mediterranean region; EUR = European region; GDP = gross domestic product; LIC = low-income; LMIC = lower middle-income; SEAR = Southeast Asian region; UMIC = upper middle-income; WPR = Western Pacific region.
